# Maternal and Neonatal Predictors of Severe Adverse Outcomes among Low-Birth-Weight Neonates in Kericho County, Kenya

**DOI:** 10.64898/2026.07.13.26357928

**Authors:** Judy Cheptoo, Vincent K. Mukthar, Morris S. Shisanya

**Author notes:** (*Corresponding Author*) |. |.

## Abstract

**Background:** Low-birth-weight (LBW) neonates carry a disproportionate share of newborn morbidity and mortality in sub-Saharan Africa. Distinguishing which maternal and neonatal characteristics mark the highest risk supports bedside risk stratification in resource-limited newborn units. This study examined maternal and neonatal predictors of severe adverse outcomes among LBW neonates admitted to a county referral hospital in Kenya.

**Methods:** A facility-based cross-sectional analysis was conducted on 169 LBW neonate–mother pairs admitted to the newborn unit at Kericho County Referral Hospital. The outcome was a severe adverse outcome, defined as a composite of respiratory distress, sepsis, hypothermia, hypoglycaemia, prolonged admission (≥ 7 days), or neonatal death. Maternal and neonatal factors were screened using chi-square, Fisher’s exact, and independent-samples t tests, with crude odds ratios (ORs) and 95% confidence intervals (CIs). A restricted multivariable logistic regression, limited to maternal and neonatal predictors, produced adjusted odds ratios (AORs).

**Results:** Severe adverse outcomes occurred in 136 of 169 neonates (80.5%). At the bivariate level, pregnancy-induced hypertension (PIH; OR = 6.69, 95% CI 1.53–29.27, p = 0.004) and preterm birth (OR = 3.79, 95% CI 1.50–9.56, p = 0.003) were associated with higher odds of a severe outcome, and severe outcomes clustered at lower birth weight and gestational age and higher neonatal risk-factor counts (all p < 0.001). In the restricted adjusted model, PIH (AOR = 7.66, 95% CI 1.56–37.55, p = 0.012) and birth weight (AOR = 0.997 per gram, p = 0.002) retained independent significance.

**Conclusion:** Neonatal biological vulnerability—particularly lower birth weight—together with maternal pregnancy-induced hypertension were the predictors most strongly and independently associated with severe adverse outcomes. Risk stratification of LBW neonates should prioritise the smallest infants and those born to mothers with hypertensive disease.

## 1. Introduction

Low birth weight (LBW), defined as a birth weight below 2,500 g regardless of gestational age, remains one of the most important markers of newborn vulnerability worldwide (1). An estimated one in seven live births, about 19.8 million infants each year, is low birth weight, and the burden is concentrated in low- and middle-income countries (2). Low-birth-weight infants may be born preterm, growth-restricted, or both, and each pathway carries a distinct but overlapping set of risks for the neonate (3).

Small and vulnerable newborns those born preterm, small-for-gestational-age, or low birth weight account for the majority of neonatal deaths globally, and preterm birth is now the leading single cause of death in children under five years (4,5). Newborn deaths have fallen more slowly than deaths later in childhood, and the neonatal period continues to concentrate risk: globally, around 2.3 million newborns die each year, most from prematurity, intrapartum-related events, and infection (6). Sub-Saharan Africa carries a disproportionate share of this burden.

In Kenya, the neonatal mortality rate has remained essentially unchanged over recent survey cycles, at approximately 21 deaths per 1,000 live births, and newborn care quality varies widely across facilities (7; 8). County referral hospitals occupy a pivotal position in the newborn care system: they receive the sickest and smallest infants, both those born on site and those referred from lower-level facilities, yet they frequently operate under constraints of staffing, equipment, and essential commodities. In such settings, the practical challenge is not only to deliver care but to recognise quickly which admitted LBW neonates are most likely to deteriorate.

Maternal and neonatal clinical characteristics available at or shortly after admission offer a pragmatic basis for such risk stratification. On the maternal side, hypertensive disorders of pregnancy, poor nutritional status, and pregnancy complications shape the intrauterine environment, and the timing of delivery, and pregnancy-induced hypertension (PIH) in particular has been linked to preterm delivery, growth restriction, and respiratory morbidity in the newborn (9). On the neonatal side, birth weight, gestational age, prematurity, and the accumulation of coexisting risk factors are consistently associated with adverse outcomes and death among LBW infants admitted for care (10; 11).

Despite this general knowledge, facility-specific evidence on which maternal and neonatal factors best distinguish severe from non-severe outcomes among admitted LBW neonates is limited in many Kenyan county referral settings. Understanding these predictors can help nurses and clinicians target monitoring, thermal care, feeding support, and early escalation to the infants who need them most. This paper therefore examines the maternal and neonatal predictors of severe adverse outcomes among LBW neonates admitted to the newborn unit at Kericho County Referral Hospital (KCRH), focusing on biological and clinical vulnerability rather than on care-process or health-system factors, which are addressed elsewhere.

## 2. Materials and Methods

### 2.1 Study design and setting

This was a facility-based analytical cross-sectional study conducted at the newborn unit of Kericho County Referral Hospital, a public referral facility serving Kericho County in the South Rift region of Kenya. The newborn unit is a high-volume setting that admits both inborn neonates and those referred from lower-level facilities within and around the county, and it manages high-risk newborns including low-birth-weight and preterm infants. The present analysis draws on the quantitative component of a larger mixed-methods study on the determinants of adverse outcomes of LBW neonates and reports maternal and neonatal predictors only; care-related, health-system, and qualitative components are reported in companion papers.

### 2.2 Study population and sample

The study population comprised low-birth-weight neonates (birth weight < 2,500 g) admitted to the newborn unit together with their mothers. Neonate–mother pairs were eligible when the neonate met the LBW definition and maternal and neonatal records contained the variables required for analysis. A total of 169 LBW neonate–mother pairs were included. Data were abstracted from maternal and neonatal records using a structured tool covering maternal socio-demographic and obstetric characteristics, neonatal clinical characteristics, and documented outcomes during admission.

### 2.3 Outcome variable

The outcome was a severe adverse neonatal outcome, defined as a composite indicator taking the value of one when a neonate experienced any of the following documented conditions during admission: respiratory distress, neonatal sepsis, hypothermia, hypoglycaemia, prolonged admission of seven days or more, or neonatal death. Because the simpler indicator of “any adverse outcome” was highly prevalent and imbalanced, the severe adverse outcome composite was used as the analytic endpoint, consistent with the parent study. Neonates not meeting any of these criteria were classified as not having a severe adverse outcome.

### 2.4 Predictor variables

Maternal predictors comprised pregnancy-induced hypertension, antenatal care attendance, maternal age, and parity. Neonatal predictors comprised birth weight (in grams), gestational age (in completed weeks), preterm birth (< 37 weeks), Apgar score, congenital anomalies, and a neonatal risk-factor count summarising the number of coexisting neonatal risk conditions present. In keeping with the scope of this paper, care-related and delay variables were deliberately excluded from the predictor set.

### 2.5 Statistical analysis

Data were analysed using standard statistical software. Categorical variables were summarised as frequencies and percentages and continuous variables as means and standard deviations.

Bivariate associations between each maternal and neonatal factor and the severe adverse outcome were tested using the Pearson chi-square test (or Fisher exact test where expected cell counts were small) for categorical predictors and the independent-samples t test for continuous predictors, with crude odds ratios and 95% confidence intervals reported for categorical factors. A restricted multivariable logistic regression model was then fitted, entering the maternal and neonatal predictors simultaneously to estimate adjusted odds ratios (AORs) with 95% confidence intervals. This model was intentionally limited to maternal and neonatal factors and is therefore narrower than the full determinants model reported in the flagship paper from this study. Model performance was assessed using the omnibus likelihood-ratio test, the Hosmer–Lemeshow goodness-of-fit test, the Nagelkerke R^2^, and the overall classification accuracy. Because the neonatal risk-factor count overlaps with its component variables (preterm birth, birth weight, Apgar score, and congenital anomalies), collinearity was examined and interpreted alongside the model estimates. A two-sided p-value below 0.05 was considered statistically significant.

### 2.6 Ethical considerations

Ethical approval was granted by the Kabarak University Research Ethics Committee (approval number KUREC-170226; reference KABU01/KUREC/001/17/02/26), and a research licence was obtained from the National Commission for Science, Technology and Innovation (NACOSTI licence number NACOSTI/P/26/4186446). Permission to access records was obtained from the hospital administration. Data were handled in accordance with the approved protocol; records were de-identified for analysis, and no individual participant is identifiable in this report.

## 3. Results

### 3.1 Sample overview

Of the 169 low-birth-weight neonates included, 136 (80.5%) experienced a severe adverse outcome, and 33 (19.5%) did not. The predominance of severe outcomes reflects the referral nature of the newborn unit and the biological fragility of the admitted LBW population. Table 1 summarises the bivariate associations of maternal and neonatal factors with the severe outcome.

**Table 1.** Bivariate associations of maternal and neonatal factors with severe adverse outcome.

| Variable | Severe<br>(n = 136) | Not Severe<br>(n = 33) | $\chi^2/t$ | Crude OR<br>(95% CI) | p |
| --- | --- | --- | --- | --- | --- |
| Pregnancy-induced hypertension (PIH) | 41 (30.1) | 2 (6.1) | 8.12 | 6.69 (1.53, 29.27) | **.004** |
| Attended ANC during pregnancy | 121 (89.0) | 30 (90.9) | 0.11 | 0.81 (0.22, 2.97) | .746 |
| Preterm birth | 122 (89.7) | 23 (69.7) | 8.73 | 3.79 (1.50, 9.56) | **.003** |

| Variable | Severe<br>(n = 136) | Not Severe<br>(n = 33) | $\chi^2/t$ | Crude<br>OR (95%<br>CI) | p |
| --- | --- | --- | --- | --- | --- |
| Congenital anomalies | 25 (18.4) | 3 (9.1) | 1.66 | 2.25 (0.64, 7.97) | .198 |
| Birth weight (g), *M* (*SD*) | 1704.8<br>(337.4) | 2113.0<br>(422.8) | 5.92 | — | <<br>**.001** |
| Gestational age (weeks), *M* (*SD*) | 31.9 (4.0) | 34.7 (3.0) | 3.83 | — | <<br>**.001** |
| Maternal age (years), *M* (*SD*) | 27.1 (6.7) | 28.3 (5.9) | 0.91 | — | .365 |
| Parity, *M* (*SD*) | 2.4 (3.2) | 2.2 (1.3) | −0.29 | — | .773 |
| Apgar score, *M* (*SD*) | 8.5 (1.5) | 8.5 (1.2) | −0.22 | — | .825 |
| Number of neonatal risk factors, *M* (*SD*) | 1.4 (0.7) | 0.9 (0.7) | −4.11 | — | <<br>**.001** |

Note. Categorical variables are n (%); continuous variables are M (SD). $\chi^2$ = Pearson chi-square; t = independent-samples t test; OR = odds ratio; CI = confidence interval; N = 169. Bold p values indicate statistical significance at $p < 0.05$ .

At the bivariate level, pregnancy-induced hypertension was present in 30.1% of neonates with a severe outcome compared with 6.1% of those without, corresponding to nearly a seven-fold increase in crude odds (OR = 6.69, 95% CI 1.53–29.27, p = 0.004). Preterm birth was likewise more common among severe cases (89.7% versus 69.7%; OR = 3.79, 95% CI 1.50–9.56, p = 0.003). Severe outcomes occurred at markedly lower mean birth weight (1,704.8 g versus 2,113.0 g) and gestational age (31.9 versus 34.7 weeks) and at a higher mean neonatal risk-factor count (1.4 versus 0.9), all differences significant at p < 0.001. Antenatal care attendance, congenital anomalies, maternal age, parity, and Apgar score were not significantly associated with the outcome.

### 3.2 Restricted multivariable model

Table 2 presents the restricted multivariable logistic regression, in which the ten maternal and neonatal predictors were entered simultaneously. After mutual adjustment, pregnancy-induced hypertension and birth weight retained independent statistical significance.

**Table 2.** Restricted Multivariable Logistic Regression of Maternal and Neonatal Predictors of Severe Adverse Outcome (N = 169).

| Predictor | Adjusted OR (95% CI) | p |
| --- | --- | --- |

| Predictor | Adjusted OR (95% CI) | *p* |
| --- | --- | --- |
| Pregnancy-induced hypertension (PIH) | 7.66 (1.56, 37.55) | .012 |
| Birth weight (g) | 0.997 (0.995, 0.999) | .002 |
| Number of neonatal risk factors | 6.93 (0.73, 65.70) | .092 |
| Preterm birth | 0.12 (0.01, 1.48) | .098 |
| Apgar score | 1.32 (0.82, 2.12) | .252 |
| Congenital anomalies | 0.39 (0.03, 5.14) | .472 |
| Gestational age (weeks) | 0.89 (0.69, 1.15) | .378 |
| Maternal age (years) | 0.98 (0.90, 1.07) | .626 |
| Parity (number of births) | 1.02 (0.88, 1.18) | .799 |
| Attended ANC during pregnancy | 0.84 (0.20, 3.58) | .809 |

**Note.** OR = odds ratio; CI = confidence interval; ANC = antenatal care. *N* = 169. The restricted multivariable logistic regression model included 10 predictors. Model fit statistics were as follows: omnibus $\chi^2(10) = 50.74$ , $p < .001$ ; Hosmer–Lemeshow $\chi^2(8) = 12.44$ , $p = .133$ ; Nagelkerke $R^2 = .413$ . The model correctly classified 85.8% of cases. The variance inflation factor for the neonatal risk-factor count was 5.16. Significant predictors ( $p < .05$ ) are shown in bold.

After adjustment for the remaining maternal and neonatal factors, pregnancy-induced hypertension was associated with a more than seven-fold increase in the odds of a severe adverse outcome (AOR = 7.66, 95% CI 1.56–37.55, p = 0.012), and each additional gram of birth weight was associated with a small but significant reduction in the odds of a severe outcome (AOR = 0.997 per gram, 95% CI 0.995–0.999, p = 0.002). Expressed per 100 g, this corresponds to roughly a 26% reduction in the odds of a severe outcome for every 100 g of additional birth weight. The neonatal risk-factor count (AOR = 6.93, p = 0.092) and preterm birth (AOR = 0.12, p = 0.098) were borderline; their wide and unstable confidence intervals reflect collinearity between the composite risk-factor count and its component variables (preterm birth, birth weight, Apgar score, and congenital anomalies), indicated by a variance inflation factor of 5.16 for the count. The model discriminated and fit the data adequately (omnibus χ^2^(10) = 50.74, p < 0.001; Hosmer–Lemeshow p = 0.133; Nagelkerke R^2^ = 0.413; 85.8% correctly classified). Figure 1 displays the adjusted odds ratios and confidence intervals.

**Figure 1.**
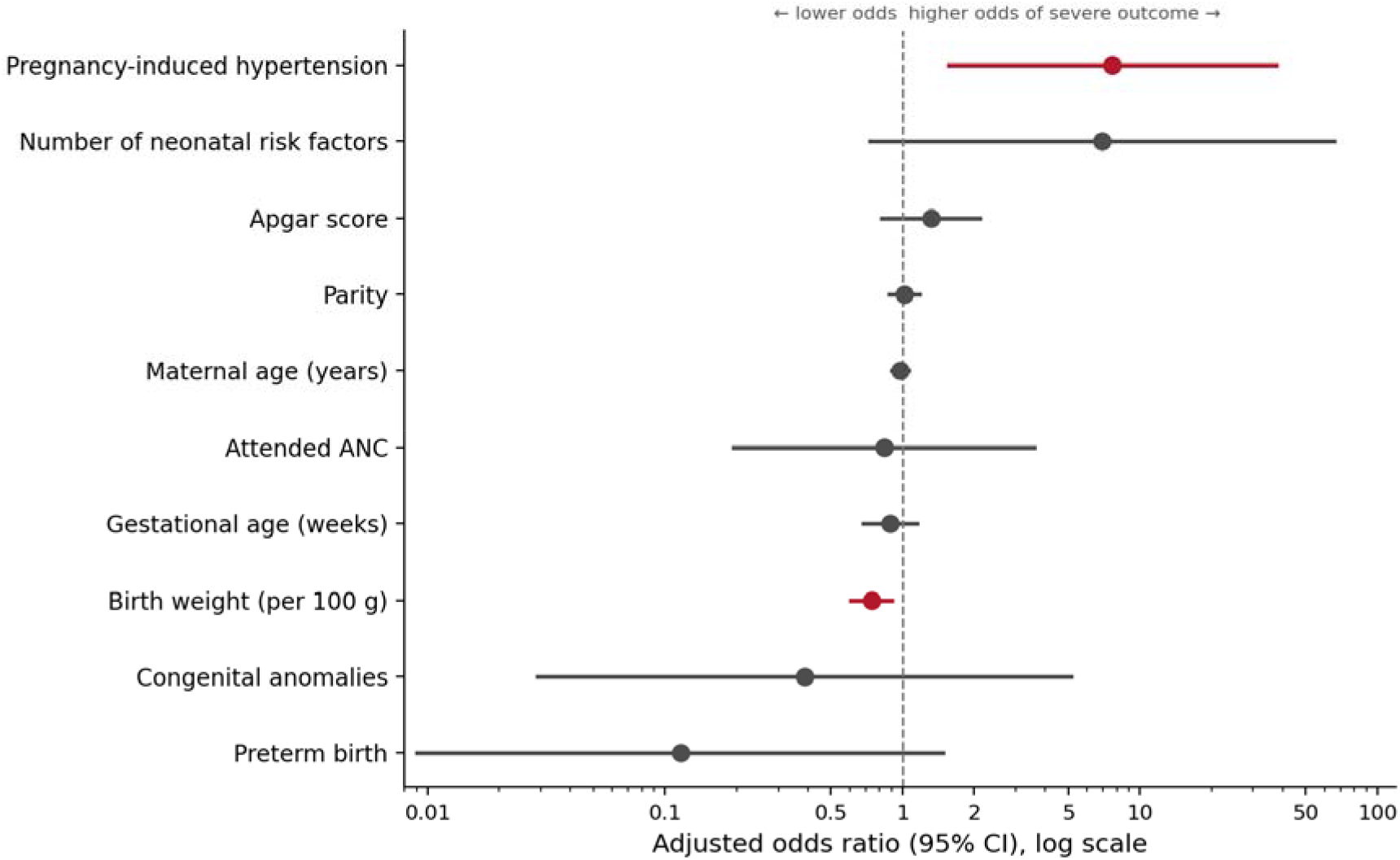
Adjusted odds ratios (95% CI) for maternal and neonatal predictors of severe adverse outcome, restricted multivariable model. *Note*. Points are adjusted odds ratios; horizontal lines are 95% confidence intervals; the dashed line marks the null value of 1. Red markers indicate predictors whose confidence interval excludes 1 (pregnancy-induced hypertension and birth weight). Birth weight is displayed per 100 g for interpretability; the model estimate in Table 2 is expressed per gram. The x-axis is on a logarithmic scale.

## 4. Discussion

This study identified two independent predictors of severe adverse outcomes among low-birth-weight neonates admitted to a Kenyan county referral hospital: maternal pregnancy-induced hypertension (PIH) and neonatal birth weight. After adjustment for potential confounders, infants born to mothers with PIH and those with lower birth weights remained significantly more likely to experience severe outcomes. The high prevalence of severe outcomes (80.5%) further highlights the substantial burden of morbidity and mortality among low-birth-weight neonates in resource-limited settings and reinforces the need for early risk identification and targeted neonatal care (4; 12).

Birth weight emerged as the strongest and most consistent predictor of adverse outcomes. Although the adjusted odds ratio per gram appeared numerically small, the cumulative clinical effect was considerable, with every 100-g increase in birth weight translating into an approximately 25% reduction in the odds of severe adverse outcomes. This finding is consistent with extensive evidence demonstrating that birth weight is one of the most powerful predictors of neonatal survival because it reflects the combined effects of prematurity, fetal growth restriction, and intrauterine health (3; 12; 13). Similar findings have been reported across sub-Saharan Africa and other low- and middle-income countries, where neonatal mortality declines progressively with increasing birth weight and gestational maturity (4; 14). From a clinical perspective, birth weight is particularly valuable because it is immediately available at admission, inexpensive to obtain, and can guide prompt allocation of intensive thermal protection, nutritional support, glucose monitoring, infection surveillance, and respiratory care (12).

Maternal pregnancy-induced hypertension independently predicted severe neonatal outcomes even after controlling for birth weight and gestational age, suggesting that its influence extends beyond its established role in causing fetal growth restriction and preterm birth. Hypertensive disorders impair uteroplacental perfusion, resulting in chronic fetal hypoxia, placental insufficiency, and altered fetal adaptation, all of which increase neonatal vulnerability immediately after birth (15; 16). Previous studies have similarly demonstrated increased risks of respiratory distress syndrome, intraventricular haemorrhage, necrotising enterocolitis, and neonatal mortality among infants born to hypertensive mothers (16; 17). The persistence of this association after multivariable adjustment indicates that maternal hypertensive disease should be recognised as an important antenatal marker for neonatal risk stratification. Strengthening communication between maternity and newborn units and ensuring that maternal obstetric diagnoses accompany neonatal referral information could facilitate earlier surveillance and intervention for these high-risk infants (12).

Prematurity and the cumulative neonatal risk-factor score showed significant associations with severe outcomes in the unadjusted analyses but lost statistical significance after adjustment. This finding most likely reflects collinearity among closely related indicators of neonatal vulnerability rather than an absence of biological effect. Birth weight, gestational age, prematurity, and composite risk scores capture overlapping dimensions of fetal maturity and physiological reserve, making it difficult to isolate their independent contributions within a relatively small dataset. Similar modelling challenges have been reported in neonatal prognostic studies, where birth weight frequently remains the dominant predictor once correlated measures of maturity are included in multivariable analyses (3; 13). Consequently, birth weight represents the most parsimonious and clinically useful bedside indicator for identifying neonates requiring enhanced monitoring.

Several maternal and neonatal characteristics, including antenatal care attendance, maternal age, parity, Apgar score, and congenital anomalies, were not independently associated with severe outcomes. The lack of association with antenatal care attendance may reflect the uniformly high coverage observed in both outcome groups, producing limited discriminatory power. Furthermore, attendance alone provides little information regarding the quality, timing, or content of antenatal services received, factors that are more closely linked to pregnancy outcomes (18). Likewise, the absence of significant associations for congenital anomalies and Apgar score should be interpreted cautiously because the relatively small number of neonates without severe outcomes reduced statistical power and may have limited the ability to detect modest effects.

The findings have important implications for neonatal care in Kenyan county referral hospitals and comparable settings across sub-Saharan Africa, where neonatal mortality remains disproportionately high and resources for specialised newborn care are constrained (6; 19). Incorporating simple admission-based risk stratification using birth weight alongside maternal hypertensive status could enable healthcare providers to prioritise intensive monitoring, optimise nurse allocation, and initiate timely supportive interventions for infants at greatest risk. Such an approach complements universal essential newborn care while promoting more efficient use of limited neonatal resources (12).

Several limitations should be considered when interpreting these findings. The cross-sectional design precludes causal inference, while the single-centre setting may limit generalisability to other health facilities with different case mixes or levels of neonatal care. In addition, the relatively small number of infants without severe outcomes resulted in an imbalanced outcome distribution, reducing statistical precision and limiting the number of predictors that could be reliably retained in the multivariable model. Nevertheless, the study has important strengths, including complete verification of the analytical dataset, use of a clinically meaningful composite measure of severe adverse outcomes, and evaluation of predictors that are readily available in routine clinical practice. These characteristics enhance the applicability of the findings and support their translation into practical risk assessment strategies for newborn care in resource-constrained settings.

## 5. Conclusion

Among low-birth-weight neonates admitted to a Kenyan county referral hospital, severe adverse outcomes were common and were most strongly and independently associated with lower birth weight and maternal pregnancy-induced hypertension. Neonatal biological vulnerability, efficiently summarised by birth weight, appears to outweigh most individually measured maternal factors after adjustment, while pregnancy-induced hypertension stands out as an upstream maternal marker that remains influential in its own right. Newborn units should use birth weight and a maternal history of hypertensive disease as immediate, low-cost triggers for closer monitoring and earlier intervention, and should strengthen the linkage of maternal and neonatal information so that the most vulnerable infants are identified from the moment of admission.

## Data Availability

All data produced in the present study are available upon reasonable request to the authors

## Declarations

## Ethical approval and consent

Ethical approval was obtained from the Kabarak University Research Ethics Committee (KUREC-170226; ref. KABU01/KUREC/001/17/02/26), and a research licence was issued by NACOSTI (NACOSTI/P/26/4186446). The study used de-identified record data under the approved protocol.

## Availability of data

The de-identified data supporting the findings of this study are available from the corresponding author on reasonable request, subject to institutional and ethical approvals.

## Funding

This research received no specific grant from any funding agency in the public, commercial, or not-for-profit sectors.

## Conflicts of interest

The authors declare that they have no competing interests.

## Authors’ contributions

JC conceived and designed the study, led data collection, and drafted the manuscript. VKM and MSS contributed to protocol conception, execution, analysis, and interpretation. Additionally, VKM revised the manuscript critically and approved the final manuscript.

## Acknowledgements

The authors thank the administration and newborn unit staff of Kericho County Referral Hospital for their support during data collection.

## Notes

### Competing Interest Statement

The authors have declared no competing interest.

### Author Declarations

1. Kabarak University Research and Ethics Committee- KUREC-170226; ref. KABU01/KUREC/001/17/02/26 2. National Commission of Science, Technology and Innovation-NACOSTI/P/26/4186446

